# Low neutralizing antibody titers against the Mu variant of SARS-CoV-2 in BNT162b2 vaccinated individuals

**DOI:** 10.1101/2021.11.19.21266552

**Authors:** Diego A. Álvarez-Díaz, Ana Luisa Muñoz, Pilar Tavera-Rodríguez, María T. Herrera-Sepúlveda, Hector Alejandro Ruiz-Moreno, Katherine Laiton-Donato, Carlos Franco-Muñoz, Dioselina Pelaez-Carvajal, Diego Cuellar, Alejandra M. Muñoz-Ramirez, Marisol Galindo, Edgar J. Arias-Ramirez, Marcela Mercado-Reyes

**Affiliations:** Grupo Genómica de Microorganismos Emergentes. Dirección de Investigación en Salud Pública, Instituto Nacional de Salud, Bogotá, Colombia; Fundación Banco Nacional de Sangre Hemolife, Bogotá, Colombia; Dirección de Investigación en Salud Pública. Instituto Nacional de Salud, Bogotá, Colombia; Grupo de Parasitología. Dirección de Investigación en Salud Pública, Instituto Nacional de Salud, Bogotá, Colombia; Dirección de Producción. Instituto Nacional de Salud, Bogotá, Colombia

**Keywords:** COVID-19, Spike protein, SARS-CoV-2 Variants, Neutralizing antibodies, Mu (B.1.621) variant, Gamma (P.1) variant

## Abstract

**Background:** Global surveillance programs for the virus that causes COVID-19 are showing the emergence of variants with mutations in the Spike protein, including the Mu variant, recently declared a Variant of Interest (VOI) by the World Health Organization. Genomic and laboratory surveillance is important in these types of variants because they may be more infectious or less susceptible to antiviral treatments and vaccine-induced antibodies.

**Objectives:** To evaluate the sensitivity of the Mu variant (B.1.621) to neutralizing antibodies induced by the BNT162b2 vaccine.

**Study design:** Three of the most predominant SARS-CoV-2 variants in Colombia during the epidemiological peaks of 2021 were isolated. Microneutralization assays were performed by incubating 120 TCDI_50_ of each SARS-CoV-2 isolate with five 2-fold serial dilutions of sera from 14 BNT162b2 vaccinated volunteers. The MN_50_ titer was calculated by the Reed-Muench formula

**Results:** The three isolated variants were Mu, a Variant of Interest (VOI), Gamma, a variant of concern (VOC), and B.1.111 that lacks genetic markers associated with greater virulence. At the end of August, the Mu and Gamma variants were widely distributed in Colombia. Mu was predominant (49%), followed by Gamma (25%). In contrast, B.1.111 became almost undetectable. The evaluation of neutralizing antibodies suggests that patients vaccinated with BNT162-2 generate neutralizing antibody titers against the Mu variant at significantly lower concentrations relative to B.1.111 and Gamma.

**Conclusions:** This study shows the importance of continuing with surveillance programs of emerging variants as well as the need to evaluate the neutralizing antibody response induced by other vaccines circulating in the country against Mu and other variants with high epidemiological impact.

**Highlights:**

- Mu and Gamma variants represented 49% and 25% of cases in Colombia by August 2021.
- Increased proportion of SARS-COV-2 cases were mostly associated with Mu variant, despite being detected simultaneously with the VOC Gamma
- The Mu variant remarkably escapes from neutralizing antibodies elicited by the BNT162b2-vaccine
- Laboratory studies of neutralizing antibodies are useful to determine the efficacy of SARS-CoV-2 vaccines against VOC and VOI.

## 1. Introduction

Between October and December 2020, genomic epidemiology data from the emerging SARS-CoV-2 lineages B.1.1.7 (Alpha), B.1.351 (Beta), and P.1 (Gamma) suggested a significant association with increased transmissibility and consequently, a global risk to public health. With this evidence, the Centers for Disease Control and Prevention (CDC) and the World Health Organization (WHO) established a hierarchical classification system to distinguish the emerging variants of SARS-CoV-2 into Variant of Interest (VOI) and Variant of Concern (VOC) [1]. To date, the WHO has designated four VOC (Alpha, Beta, Gamma, and Delta) and five VOI (Eta, Iota, Kappa, Lamda, and Mu). However, a VOI might escalate to VOC if, besides the presence of genetic markers associated with higher virulence supported with epidemiological data suggesting it as an emerging risk to global public health, because there is solid evidence of negative impacts in public health including increased transmissibility, virulence, decreased effectiveness of therapeutic measures, vaccines or diagnostics [2].

The new SARS-CoV-2, B.1.621 lineage detected in January 2021, was proposed as a variant of interest (VOI) following the SARS-CoV-2 genomic surveillance in Colombia between December 2020 and April 2021 [3]. By the end of August 2021, the WHO confirmed the VOI status of this lineage and assigned it the “Mu” letter of the Greek alphabet [2]. Since its detection, the Mu variant has spread to 43 countries, with a higher presence in the British Virgin Islands, Colombia, Dominican Republic, Ecuador, and Haití [4]. In Colombia, B.1.621 spread from the Caribbean region to the rest of the country within six months, and by the end of August, was the predominant lineage [5].

Preliminary studies assessing the sensitivity of the Mu variant to antibodies induced by the BNT162b2 vaccination using pseudoviruses and replication-competent SARS-CoV-2 yielded contradictory results [6,7]. In this study, we determined the neutralizing antibody titers in BNT162b2-vaccinated individuals against SARS-CoV-2 isolates from the Mu, Gamma, and B.1.111 lineage, using microneutralization assays.

## 2. Materials and methods

The spatiotemporal distribution of SARS-CoV-2 lineages circulating in Colombia between January and August 2021 was determined following the National Program for the Genomic Characterization of SARS-CoV-2 based on representativeness and virologic criteria for probabilistic sampling [3,8]. Then, we isolated the three most predominant SARS-CoV-2 lineages during this period, used to evaluate the titer of neutralizing antibodies by microneutralization assays (MN) as previously described [9], briefly Two-fold serial dilutions ranging from 1:4 to 1:2460 of serum samples were incubated with 120 TCDI_50_ of each variant. We tested sera from 14 volunteers, collected between 72 and 87 days after receiving the second dose of the BNT162b2 COVID-19 vaccine. Individuals with previous or current SARS-CoV-2 infection or during clinical follow-up, or the presence of total antibodies against SARS-CoV-2 at the time of the first dose of the vaccine administration, were excluded. The protective neutralizing antibody titer that prevented cytopathic effect in 50% of the wells (MN_50_) was calculated by the method of Reed and Muench [10]. All neutralization assays with infectious SARS-CoV-2 viruses were conducted in a biocontainment laboratory.

Sera were first screened for the absence of IgG anti-nucleoprotein antibodies using a qualitative ELISA (ID Screen SARS-CoV-2-N IgG Indirect, ID Vet). subsequently, to determine the concentration of anti-Spike IgG antibodies, samples were tested with the SARS-CoV-2 IgG assay (sCOVG) on the ADVIA Centaur XPT platform (Siemens) using the kit cut-off value (reactive ≥1.0 U/mL); the results were expressed in binding antibody units per milliliter (BAU)/mL using the conversion factor of 21.8 determined by the manufacturer based on WHO first standard 20/136 [11].

## 3. Results

### Mu and Gamma variants dominated the second epidemic peak of SARS-CoV-2 in Colombia

Three SARS-CoV-2 variants were isolated, Mu (B.1.621 lineage, GISAID ID EPI_ISL_1821065), Gamma (P.1 lineage, GISAID ID EPI_ISL_2500971) classified as a variant of concern (VOC) by the WHO and B.1.111 (GISAID ID EPI_ISL_526971) that lacks genetic markers associated with greater virulence. Their profile of mutations in S and other regions is shown in table 1. Molecular epidemiologic data allowed us to determine the spatiotemporal distribution of the most representative SARS-CoV-2 lineages in Colombia during the two epidemiological peaks of SARS-CoV-2 in 2021. The sequences obtained by probabilistic sampling indicated a significant increase of Mu (B.1.621) and Gamma (P.1) which were detected during the second peak in opposite regions of the country by the end of December 2020. While Mu was identified in the Caribbean region at the north, Gamma was identified in the Amazon region at the south. Then, by the end of August 2021, those lineages were dominant and widely distributed across the country, being the majority (74%) of COVID-19 cases in Colombia. Remarkably, Mu represented almost twice the sequences of Gamma and half of the total sequences of SARS-CoV-2 (49% of cases) in the country by the end of the third peak. In contrast, B.1.111 was distributed at the country level and dominated the first epidemic peak, then became almost undetectable by the end of the third peak (Fig. 1).

**Table 1.** Genomic characteristics of the viral isolates selected for MN_50_ assays.

| **Pango Lineage\***<br>**(WHO status)** | **Isolate name**<br>**(GISAID ID.)** | **AA Substitutions** |
| --- | --- | --- |
| B.1.621 (Mu) | EPI_ISL_1821065 | Spike D614G, Spike D950N, Spike E484K, Spike ins145N, Spike N501Y, Spike P681H, Spike R346K, Spike T95I, Spike Y144T, Spike Y145S, N T205I, NS3 Q57H, NS3 V256I, NS8 P38S, NS8 Q72R, NS8 S67F, NS8 T11K, NSP3 A562T, NSP3 T237A, NSP3 T720I, NSP4 T492I, NSP6 Q160R, NSP12 P323L, NSP13 P419S |
| P.1 (Gamma) | EPI_ISL_2500971 | Spike D138Y, Spike D614G, Spike E484K, Spike H655Y, Spike K417T, Spike L18F, Spike N501Y, Spike P26S, Spike R190S, Spike T20N, Spike T1027I, Spike V1176F, N D415G, N G204R, N P80R, N R203K, NS3 S253P, NS8 E92K, NSP1 P80L, NSP3 K977Q, NSP3 S370L, NSP3 T1303I, NSP3 V1253F, NSP6 F108del, NSP6 G107del, NSP6 S106del, NSP13 E341D |
| B.1.111 | EPI_ISL_526971 | Spike D614G, N M234I, NS3 Q57H, NSP3 S1285F, NSP12 P323L |

\*Pango v.3.1.11 2021-08-24

**Figure 1.**
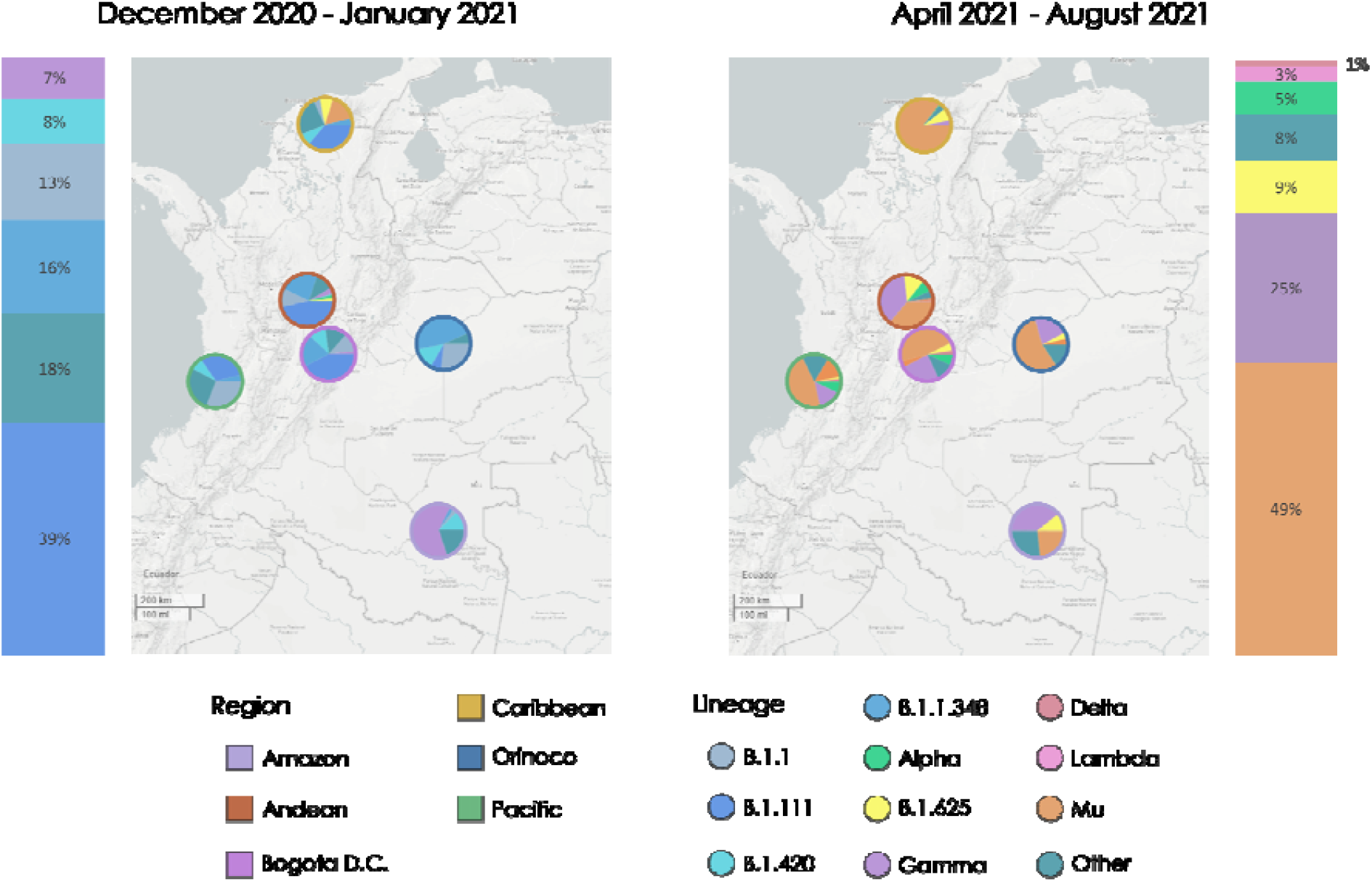
Spatiotemporal distribution of the most representative SARS-CoV-2 lineages in Colombia (bars) and its regions (map). SARS-CoV-2 lineage distribution in the five Colombian regions and Bogotá D.C December 2020 - January 2021 (left), and April-August 2021 (right). Ring colors represent the region. Interactive map available at https://microreact.org/project/6GjGXeoUW7uVauMTFCFEkE/d9357c6c [12]

### Mu and Gamma SARS-CoV-2 variants escape from neutralization by BNT162b2-vaccine serum samples

A panel of human sera from 14 volunteers vaccinated with BNT162b2 (age range, 27-58 years) was evaluated. Anti-Spike IgG antibody titers but no anti-nucleoprotein IgG antibodies were identified in all samples, suggesting the exclusive presence of vaccination-induced antibodies.

Serum samples exhibited robust neutralization against the B.1.111 lineage, with a geometric mean titer (GMT) of 224.2 TCID_50_ (Fig. 2). For the Gamma variant, the GMT was 65.2 TCID_50_, which, was 3.4 times lower compared to B.1.111.

**Figure 2.**
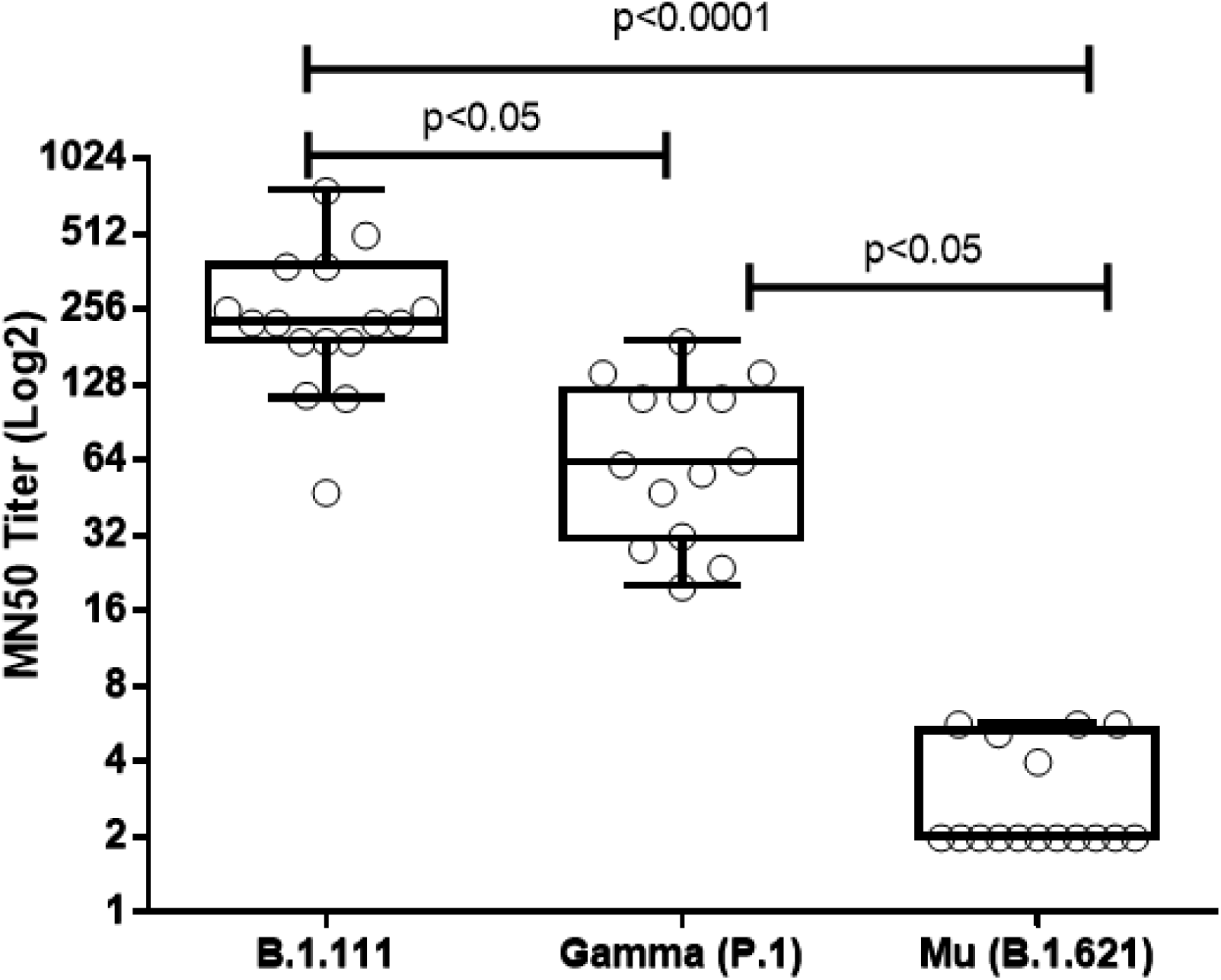
Neutralizing titers of BNT162b2 vaccinated volunteers against Mu, Gamma, and B.1.111 SARS-Cov-2 isolates. TCID_50_-based assays were performed by incubating 120 TCDI_50_ of each SARS-CoV-2 isolate with five 2-fold serial dilutions of sera from BNT162b2 vaccinated volunteers. The MN_50_ titer was calculated by the Reed-Muench formula. Statistical differences between the median values of MN_50_ titers against Mu, Gamma, and B.1.111 variants were determined using the Kruskal-Wallis test followed by Dunn’s post hoc test for multiple comparisons. An arbitrary MN_50_ titer value of 2 was assigned to the 11 out of 16 serum samples that do not neutralize the virus at the lowest dilution (<1/4).

In contrast, the GMT of 2 TCID_50_ determined for the Mu variant, was 41 and 20 times significantly lower compared to the B.1.111 and Gamma lineages, respectively (P <0.0001). It is important to note that 11 out of 14 serum samples (78.5 %) did not neutralize the virus at the dilution 1:4, which was the lowest evaluated (Fig. 2).

Finally, a strong correlation was observed between the neutralizing antibody titers (MN_50_) and IgG anti-S antibodies (BAU/mL) for B.1.111 compared with Mu. In contrast, no significant correlation was observed for Gamma (Table 2).

**Table 2.** Comparison of MN_50_ titers and binding antibody units.

| Comparison |  | Spearman r | p value* | 95% Confidence Interval |
| --- | --- | --- | --- | --- |
| MN50 B.1.111 | Anti-S IgG titer BAU/mL | 0,6861 | 0,0042 | 0,2735 to 0,8854 |
| MN50 Gamma (P.1) | Anti-S IgG titer BAU/mL | 0.4619 | 0,0978 | -0,1084 to 0,8035 |
| MN50 Mu (B.1.621) | Anti-S IgG titer BAU/mL | 0,5387 | 0,0338 | 0,04256 to 0,8218 |

\*Two-tailed

## 4. Discussion

Molecular epidemiologic dynamics of SARS-CoV-2 lineages in Colombia during the second and third epidemiological peaks between January and August 2021 suggest a significant community transmission of the emerging variants Mu (B.1.621) and Gamma (P.1) which led to the rapid dispersal from the Caribbean and Amazon region and displacement of the dominant lineages by August 2021. Furthermore, according to the INS epidemiological surveillance system, the number of confirmed cases of SARS-CoV-2 during the second epidemiological peak was almost double that reported during the first peak [13]. Hence, as the Mu and Gamma variants represented 49% and 25% of the SARS-CoV-2 sequences respectively, it is probable that these variants are associated with the increase in the number of cases of the third epidemiological peak in Colombia, although the greater representativeness of Mu potentially implies a greater epidemiological impact of this variant (Fig. 1).

In this study, a 3.4-fold decrease in the neutralization of Gamma relative to B.1.111 variant was observed (Fig. 2); similar results were found in previous studies reporting 3 to 5.12-fold reduced sensitivity of variant Gamma to serum from individuals vaccinated with the Pfizer vaccine BNT162b2 using pseudovirus bearing the Gamma or wild-type SARS-CoV-2 S protein, with D614G exchange [14,15].

On the other hand, the sera from BNT162b2 vaccinated volunteers exhibited a robust decrease in the neutralization of Mu by 43.2 and 12.56-fold, relative to B.1.111 and Gamma, respectively (Fig. 2). While this was consistent with the report by Uriu et al. who report lower neutralization titers against Mu relative to a parental D614G variant and Gamma [15], the magnitude of the difference was more noticeable in the present study. By contrast, Messali et al. reported a slightly lower neutralization titer in the sera from BNT162b2 vaccinated volunteers against Mu relative to a B.1 isolate. Although the differences observed between these studies may be due to the use of different platforms for the screening of neutralizing antibodies (i.e pseudovirus or isolates), all these evidenced the resistance of the Mu variant to antibodies elicited by the BNT162b2 vaccine.

Remarkably, the genomic surveillance and laboratory studies with the emerging lineage B1+L249S+E484K identified simultaneously with the Mu variant in the same geographic region of Colombia, evidenced reduced neutralization of convalescent sera accompanied by a decline of cases associated with this variant, demonstrating that this lineage does not represent a concern for public health in Colombia [16][9]. Therefore, the escape of vaccine antibodies and the dramatic increase in cases associated with Mu in Colombia, even with a greater impact than the VOC Gamma, suggest that Mu can be classified as VOC, depending on the dispersion and global cases in the incoming months. Nonetheless, further studies evaluating the cell responses to SARS-CoV-2 vaccination must be included to better assess the immunologic effects of SARS-CoV-2 vaccines.

Finally, this study shows the importance of continuing with the surveillance programs of emerging variants, as well as the need to evaluate the resistance of VOC/VOIs to humoral immunity elicited by the vaccines circulating in the country.

## Data Availability

All data produced in the present study are available upon reasonable request to the authors

## Ethics

The study protocol was approved by the Ethics Committee of the Colombian National Health Institute, (CEMIN)-04-2021, carried out in accordance with the Declaration of Helsinki. All subjects enrolled in this research responded voluntarily to an informed consent formulary.

## Acknowledgements

The authors thank the study participants for their dedication, the National Laboratory Network for routine virologic surveillance of SARS-CoV-2 in Colombia. We also thank all researchers who deposited genomes in GISAID’s EpiCoV Database contributing to genomic diversity and phylogenetic relationship of SARS-CoV-2.

